# ToKSA - Tokenized Key Sentence Annotation - a Novel Method for Rapid Approximation of Ground Truth for Natural Language Processing

**DOI:** 10.1101/2021.10.06.21264629

**Authors:** Cameron J Fairfield, William A Cambridge, Lydia Cullen, Thomas M Drake, Stephen R Knight, Neil Masson, Nicholas L Mills, Riinu Pius, Catherine A Shaw, Honghan Wu, Stephen J Wigmore, Athina Spiliopoulou, Ewen M Harrison

**Affiliations:** Centre for Medical Informatics, Usher Institute, University of Edinburgh, Edinburgh, United Kingdom; Department of Clinical Surgery, Division of Health Sciences, University of Edinburgh, Edinburgh, United Kingdom; Department of Radiology, NHS Lothian, Edinburgh, United Kingdom; Centre for Cardiovascular Science, Queen’s Medical Research Institute, University of Edinburgh, Edinburgh, United Kingdom; Institute of Health Informatics, University College London, London, United Kingdom; Centre for Population Health Sciences, Usher Institute, University of Edinburgh, Edinburgh, United Kingdom

**Keywords:** Natural language processing, Radiology Information Systems, Machine Learning, Data Curation, Electronic Health Record

## Abstract

**Objective:** Identifying phenotypes and pathology from free text is an essential task for clinical work and research. Natural language processing (NLP) is a key tool for processing free text at scale. Developing and validating NLP models requires labelled data. Labels are generated through time-consuming and repetitive manual annotation and are hard to obtain for sensitive clinical data. The objective of this paper is to describe a novel approach for annotating radiology reports.

**Materials and Methods:** We implemented tokenized key sentence-specific annotation (ToKSA) for annotating clinical data. We demonstrate ToKSA using 180,050 abdominal ultrasound reports with labels generated for symptom status, gallstone status and cholecystectomy status. Firstly, individual sentences are grouped together into a term-frequency matrix. Annotation of key (i.e. the most frequently occurring) sentences is then used to generate labels for multiple reports simultaneously. We compared ToKSA-derived labels to those generated by annotating full reports. We used ToKSA-derived labels to train a document classifier using convolutional neural networks. We compared performance of the classifier to a separate classifier trained on labels based on the full reports.

**Results:** By annotating only 2,000 frequent sentences, we were able to generate labels for symptom status for 70,000 reports (accuracy 98.4%), gallstone status for 85,177 reports (accuracy 99.2%) and cholecystectomy status for 85,177 reports (accuracy 100%). The accuracy of the document classifier trained on ToKSA labels was similar (0.1-1.1% more accurate) to the document classifier trained on full report labels.

**Conclusion:** ToKSA offers an accurate and efficient method for annotating free text clinical data.

## BACKGROUND AND SIGNIFICANCE

Natural language processing (NLP) is one of the fastest growing techniques to convert electronic health record entries into meaningful research-optimized data[1]. Unstructured free-text health records are optimized for clinical workflow and there are no established methods that can consistently extract data on disease-status from such text. At present many clinical studies rely on disease codes recorded in national registries. These registries typically record codes for disease considered to contribute to a hospital episode. However, it is likely that many conditions of interest are identified throughout the course of a healthcare episode but are not recorded in these registries. Radiology reports comprise a large resource of free text likely to contain such conditions but conversion of this free text into discrete labels is a time-consuming task[2,3].

Annotating free text reports with discrete labels can take several minutes for a single report and may take several months of full-time work for large datasets. Generating these labels can be performed manually (each report is read by a human rater who assigns diagnostic codes or other labels) or automatically (a rules-based or machine-learning-based NLP technique automatically assigns labels to reports). The latter techniques usually rely on a subset of reports which have been annotated manually to allow supervised learning[4–6]. Labels generated manually are often referred to as the “ground truth”. Annotation is repetitive and it is likely that similar or identical sentences will be encountered in multiple reports, during which clinician raters will follow similar patterns of deductive reasoning. Some of these sentences may be sufficient to annotate the report without reading the remainder of the report, and by doing so create an approximation of ground truth. By prioritizing sentences that occur across multiple reports, a label can be generated for multiple reports simultaneously. This process can further be enhanced by using disease-specific glossaries to prioritize sentences most likely to allow adequate annotation and through deduplication of similar sentences. To realize this idea, this paper proposes an approach called Tokenized Key Sentence Annotation (ToKSA). ToKSA is likely to save considerable effort for clinicians and researchers in annotating radiology reports with ground truth labels. A flowchart of the traditional approach of annotating versus the ToKSA approach is shown in Figure 1.

**Figure 1:**
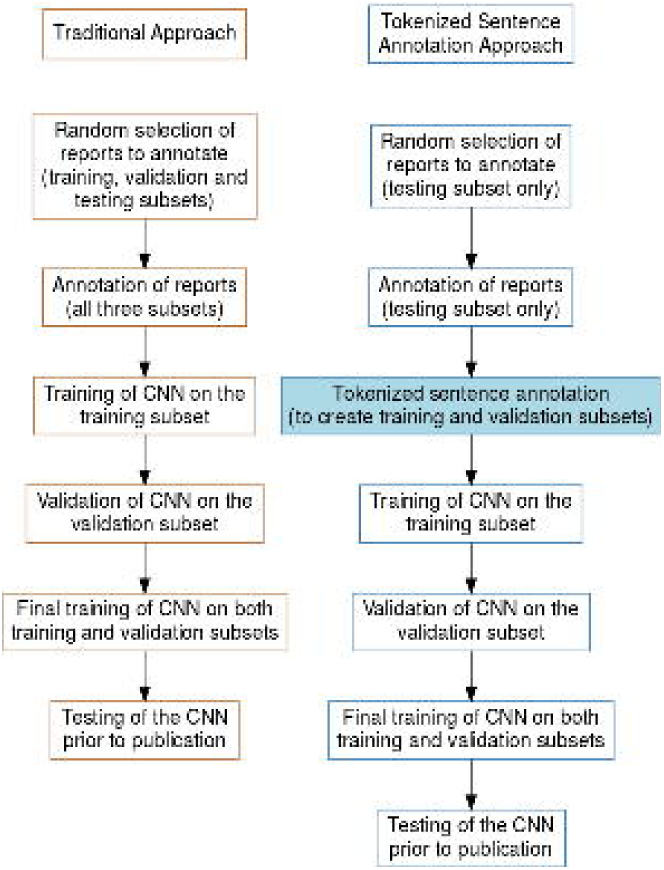
Flowchart showing typical annotation tasks to generate labels for a corpus or radiology text. The right-hand side shows the suggested strategy using Tokenized Key Sentence Annotation.

In this paper we apply ToKSA to a task where abdominal ultrasound scan (AUSS) reports are to be annotated with three labels: 1. Symptoms suspicious or not suspicious for gallstone disease (yes/no), 2. Gallstone status (present/absent/not documented) and 3. Cholecystectomy status (evidence of cholecystectomy/no evidence of cholecystectomy). Gallstones were chosen as they are common, frequently detected incidentally, and often described whether present or absent. Gallstone disease is present in around 15% of the adult population and contributes to significant morbidity and healthcare expenditure[7– 10]. As gallstones frequently occur as incidental findings[9], automatic detection of gallstones from radiology reports represents an important opportunity to supplement healthcare registries.

## OBJECTIVE

The primary aim is to assess the accuracy of Tokenized Key Sentence Annotation (ToKSA), compared to annotation of full reports, in deriving an approximation of ground truth for a document classification task. The secondary aim is to evaluate the utility of ToKSA in developing NLP models. We compare the accuracy of a convolutional neural network (CNN) trained on ToKSA-derived labels to a CNN trained on labels derived from annotation of full reports.

## MATERIALS AND METHODS

### Report Annotation

Reports from all AUSS performed for any indication between 2007 and 2018 in a tertiary health board were retrieved and anonymized. The health board covers a population of over 850,000 individuals. AUSS were undertaken across 8 separate institutions within the health board covering both inpatient and outpatient investigations. In total 180,050 reports were available for analysis. Full reports were first split into “indication” and “findings” sections using regular expressions (regex)[11].

The reports were passed through cleaning and simplification (this was conducted twice, firstly for the “indication” and secondly for the “findings” sections; See Supplementary Materials for explanations and examples for specific steps):

1. Text was converted to lower case
2. The “indication” and “findings” sections were tokenized into single words and a term-frequency matrix of the words was generated
3. A term-specific glossary (for the “indication” section: symptoms relevant to the disease and disease terms; for the “findings” section: disease terms and anatomic structures affected by the disease) was created and supplemented with regex to account for variations in spelling, punctuation and word-spacing
4. Spelling mistakes were accounted for using the Damerau-Levenshtein distance (edit distance incorporating typographical errors including substitutions, insertions, deletions and transpositions)[12–14]
  a. Each word in the term-frequency matrix was joined with any terms of interest identified using an edit distance of 2 or less (1 for words with a string length of 4 or less)
5. Genuinely misspelled terms of interest were overwritten with the correct spelling after manual inspection
6. Further searching with regex was conducted to identify and overwrite misspellings with an edit distance greater than 2
7. Regex was used to overwrite patterns that are not relevant to the research question and increase the number of unique sentences
  a. Patterns included structured numeric values (e.g. patient identification number, telephone number), email address, dates, years and measurements (e.g. “12mm”)
  b. Words were overwritten by the type of matched pattern (e.g. “2007” was converted to “[year]”)
8. A new term-frequency matrix was generated encompassing word substitutions from steps 5-7
9. Each term occurring 5 or fewer times was overwritten with “[redacted]”
10. The reports were then tokenized into whole sentence N-grams
11. The sentences were arranged into a term-frequency matrix and sorted based on the frequency of the occurrence of the sentence
12. Two groups of sentences were taken forward for annotation: the 1000 most frequent sentences which contained a term of interest and the 1000 most frequent sentences without a term of interest (the latter was inspected to ensure no terms of interest were missed from the term-specific glossary).
13. The sentences were annotated with desired labels
14. The labels generated for each sentence were combined into full report labels (See Supplementary Material for further explanation of label combining)
15. Any conflicting labels were resolved through inspection of the full report

Specific examples of the regex used for annotating the AUSS reports are provided in Supplementary Material.

Each sentence was annotated independently by two clinician raters (clinician or medical student) with disagreements resolved by a third rater. Interobserver variability was calculated using a weighted Cohen’s *к* [15] and weighted Bangdiwala’s B-statistic[16]. The B-statistic corrects for agreement that arises through chance alone and maintains stable performance with marginal distributions seen in imbalanced data.

In theory, no reports should receive contradictory classifications (e.g. both “definite gallstones” and “no gallstones”), such dual classifications were sought and if present assessed for the reason underlying this.

### Evaluation of Technique Performance

To assess performance of ToKSA compared to annotation of full reports, a cohort of 3,000 randomly selected full reports were annotated. During refinement of our approach a further 407 reports were annotated giving 3,407. Each full report was also annotated by two clinician raters with disagreements resolved by a third rater. Clinician raters were blinded to labels generated by ToKSA. If one of the annotated full reports was identical to another report in the corpus, the other report was also given the same label. Agreement between ToKSA and annotation of the full reports was calculated using a weighted Cohen’s *к* [15] and weighted Bangdiwala’s B-test[16]. Variability between the techniques was shown through Bangdiwala interobserver agreement chart[17–19].

To assess the efficiency of the technique we compared anticipated reading times to read all of the reports labelled by ToKSA. We based our analysis on a published meta-analysis of average words per minute read in English in which the average silent reading time was found to be 239 words per minute[20] and calculated the reading time for the full reports and then for the 2,000 key sentences.

### Convolutional Neural Networks for Document Classification

Derivation of labels through manual annotation is usually followed by an automated classification technique such as a CNN to generate labels for the remaining reports. Whilst ToKSA generates labels for multiple reports, the most frequent sentences will not cover every report meaning an automated technique is still required. Maximizing the number of reports with labels through ToKSA may improve CNN performance by enabling a larger training set for supervised learning but may also result in decreased CNN performance for reports not containing any frequent sentences. Therefore, the performance of CNNs trained on the ToKSA-derived labels was compared to CNNs trained on labels from full reports. Two separate CNNs per annotation task were developed. One was trained on 60% (training set) and validated on 20% (validation set) of reports with a label assigned during annotation of full reports and tested on the remaining 20% (testing set) immediately prior to publication. The second was trained and validated on the reports with a label assigned by ToKSA and then tested on the same testing set as the first CNN (reports in the testing set were excluded from the training and validation set). Two CNNs were developed for each of the labels generated (symptom status, gallstone status and cholecystectomy status) resulting in 6 separate CNNs. The CNNs[21] were developed with Keras[22] and a Tensorflow[23] backend.

Text was converted into a dense matrix with each word represented by its frequency ranking. Each CNN was trained on its training set and performance monitored using the validation set. A small number of hidden layers and epochs were used initially and iteratively increased to optimize performance. The maximum length was set to the longest report such that all the data was considered, layers were set to a dropout rate of 0.15 to prevent over-fitting with a “tanh” activation of nodes in early layers and “sigmoid” activation in later layers. The loss function was optimized for binary cross entropy for symptom status and cholecystectomy status and for categorical cross entropy for gallstone status (gallstone status had three categories as “Unknown” was necessary). The difference in correctly classified reports between the two CNNs was assessed (correct classification was tested in the same testing set for all analyses). To ensure performance of the CNN trained on ToKSA-derived labels remained high for reports which lacked frequent sentences (and hence did not receive a label from ToKSA), the accuracy of the CNN classifications was also compared against reports which only received a label from the annotation of full reports.

The script for performing ToKSA is available at: https://github.com/SurgicalInformatics/ToKSA.git.

Ethical approval was granted by Lothian NHS Board South East Scotland Research Ethics Committee 01 (REC reference number 21/SS/0003). All data were deidentified and the need for individual consent was waived.

## RESULTS

In total, 180,050 AUSS reports for 116,591 individuals were available. The age of patients ranged from 16-89. 69,583 (59.7%) were female, 45,861 (39.3%) and 1,147 (1.0%) did not have sex recorded.

### Tokenization of Reports

The results of tokenization, word substitution and conversion of sentence tokens into a term-frequency matrix is shown in Supplementary Figure 1. After tokenization, the “indication” section, 385,463 unique sentences were available for annotation. After tokenization, the “findings” section, 521,524 unique sentences were available to annotate. The top 20 most frequent sentences with a term of interest in each category are shown in Supplementary Tables 1-2.

### Report Annotation

ToKSA resulted in labels for 70,000 “indications” and 85,177 “findings” sections. The remaining reports had no sentences within the 2,000 selected sentences or a label could not be derived based on annotation of the 2,000 sentences. After applying labels to the 3,407 manually annotated reports and applying these labels to any other identical reports, labels based on full report annotation were available for 10,910 “indications” and 9,286 “findings” sections. Annotation of single sentences was much faster and less repetitive than for full reports and resulted in a label being generated for more than 6 times as many reports. A total of 27 reports out of the 85,177 annotated (for the “findings” section) received conflicting annotations using ToKSA. On closer inspection of the full reports all 27 were found to contain contradictory statements. For example: “Multiple small gallstones seen in the gallbladder. No gallbladder polyps, lesions or stones.”. These reports were all annotated as “Gallstones” on the basis that the latter sentence may be routinely dictated for several reports and may have been inadvertently typed or dictated whilst the sentence confirming stones is unlikely to have been documented by mistake.

### Interobserver Agreement

Interobserver agreement between clinician raters was very high. There were 6 interobserver agreement rates assessed for clinician raters. For the “indications” section of the reports there was one label (“Suspicious” or “Not Suspicious” for gallstones), and for the “findings” section two labels were generated (1. presence or absence of gallstones and 2. presence or absence of cholecystectomy). These three measures of agreement were repeated for both the traditional full-report-based annotations (3,407 full reports annotated by 2 clinician raters) and for ToKSA (2,000 of the most common sentences annotated by 2 clinician raters). Weighted Cohen’s Kappa ranged from 0.956 to 1 and Bangdiwala’s B-statistic from 0.965 to 1 for agreement between clinician raters (Supplementary Table 3).

### Evaluation of ToKSA Performance

The concordance between ToKSA-derived labels and labels derived from annotation of the full reports was very high. Of the 1,813 reports in which both approaches generated a label for the “indication”, the misclassification rate of ToKSA was 1.6%. Weighted Cohen’s kappa was 0.927 and weighted Bangdiwala’s B-statistic was 0.98. Of 224 “indications” considered “not suspicious” for gallstones, 12 were incorrectly annotated by ToKSA as “suspicious”. Of 1,589 “indications” considered as “suspicious” for gallstones, 17 were incorrectly annotated as “not suspicious”. Of the 2,047 reports in which a label for gallstone status was generated by both ToKSA and through annotation of full reports, only 16 (0.8%) were misclassified by ToKSA. Two were incorrectly annotated by ToKSA as “No Gallstones” when gallstones were observed and 14 were incorrectly annotated as “Unknown” by ToKSA instead of “No Gallstones”. Weighted Cohen’s kappa was 0.971 and weighted Bangdiwala’s B-statistic was 0.991. Of the 1,673 in which both approaches generated a label for cholecystectomy, 100% were given the same label meaning Weighted Cohen’s kappa and weighted Bangdiwala’s B-statistic were both 1. Agreement between the techniques for the classifications made (gallstone status, cholecystectomy status and indication suspicious for gallstones) is shown in Figure 2. Confusion matrices of the performance of ToKSA versus full report annotation are shown in Supplementary Tables 4-6.

**Figure 2:**
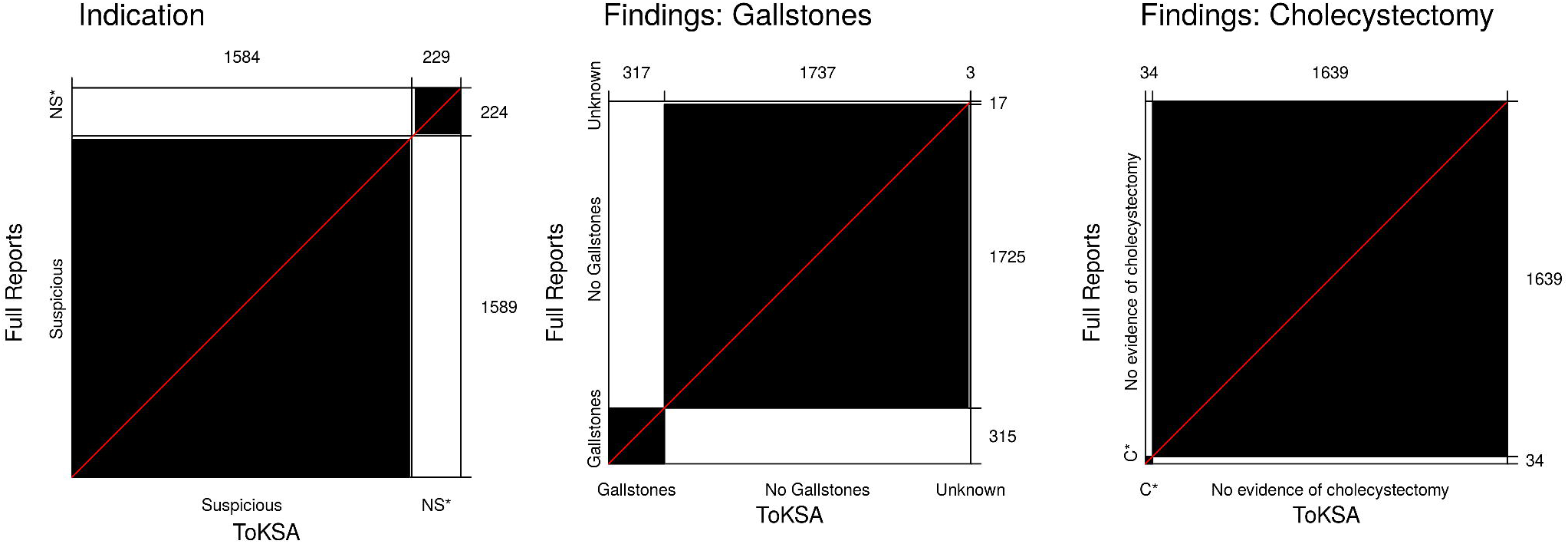
Bangdiwala agreement charts. The extent of agreement between annotation of the full AUSS reports and annotation of specific sentences within the reports is represented by the size of the black boxes inside grey rectangles located in the diagonal of the plot. The larger the black area, the greater the extent of agreement. The shaded grey rectangles represent the maximum possible agreement, given the total number of annotations. The x axis represents the number of individual reports annotated with each classification by the sentence-specific approach and the y-axis the annotations for full report approach. C*: Cholecystectomy

The indications section of the reports annotated for symptom status contained a total of 1,138,704 words (request length ranged from 1 to 540 words) with an estimated reading time of 79.7 hours. When considering only the 2,000 sentences necessary to generate labels for the same reports, a total of 6,440 words were present with an estimated reading time of 27 minutes. For the determination of gallstone status, 4,992,590 words (report length ranged from 2 to 442 words) were present in all reports for which a label was generated by ToKSA with an estimated reading time of 349.6 hours. The number of words in the 2,000 key sentences necessary to generate labels for these approaches is 12,434 with an estimated reading time of 52 minutes. For the determination of cholecystectomy status, 4,443,000 words were present in the full reports with an estimated reading time of 311.1 hours. The same 2,000 sentences were used as for gallstone status (12,234 words with 52 minutes of estimated reading time). Whilst these differences represent between a 176- to a 401-fold reduction in reading time this does not account for time taken to actually type a label and does not account for time taken during the initial setup of ToKSA.

### Convolutional Neural Network

When training the CNN on data annotated using ToKSA, the accuracy of the CNN was higher than that trained on data annotated from only the full reports. The accuracy when using a ToKSA-trained CNN compared to the full-report-trained CNN was 98.9% versus 98.3% (25 versus 38 of 2182 reports misclassified; Bangdiwala’s B-statistic 0.977 versus 0.966) for the “indication” section, 99.7% versus 98.6% (16 versus 26 of 1857 reports misclassified; Bangdiwala’s B-statistic 0.996 versus 0.996) for the determination of gallstone status and 100% versus 99.9% (0 versus 1 of 1857 reports misclassified; Bangdiwala’s B-statistic 1 versus 0.999) for cholecystectomy. A total of 111,280 (61.8%) scans were classified as symptoms suspicious for gallstones with 68,770 (38.2%) as not suspicious. 34,699 (19.3%) showed evidence of gallstones, 101,979 (56.6%) showed evidence of the absence of stones and 43,372 (24.1%) did not provide evidence on gallstone status. 10,277 (5.7%) showed evidence of cholecystectomy and 169,773 (94.3%) did not describe evidence of cholecystectomy.

Accuracy of the CNN trained on ToKSA data was similar when limited to reports which only received a label from annotation of the full reports. For the “indication” section 1793/1824 (98.3%) were correctly classified, for gallstone status 1405/1428 (98.3%) were correctly classified, and for cholecystectomy status 1505/1505 (100%) were correctly classified.

## DISCUSSION

We demonstrate a novel approach for rapid annotation of radiology reports based on Tokenized Key Sentence Annotation. ToKSA achieved high levels of accuracy when compared to conventional annotation of the full reports and offers far greater efficiency for large datasets. We demonstrate that by annotating a selection of 2,000 simplified sentences that occur frequently across all reports ToKSA was able to annotate over 85,000 reports covering almost half of the dataset. We demonstrate substantial reductions in anticpated reading time to generate these labels and we demonstrate that ToKSA can be adapted to three separate document classification tasks through modification of the term-specific glossary. Finally, we demonstrate that CNN-based approaches may offer superior performance when trained through an expanded corpus of annotated text generated by ToKSA rather than a smaller subset in which full reports are annotated.

Overall, our approach demonstrated very high levels of accuracy with only 16 (0.8%) misclassifications for gallstones and no misclassifications for cholecystectomy. For the indications section the rate of misclassification was slightly higher at (1.6%) which is most likely due to the unstructured free text entered to the scan request. In our health board, radiology reports are generally dictated with each radiologist relying on relatively systematic and standardized sentence structures and statements in each report. The indications section generally has more spelling mistakes, greater variability in use of punctuation, greater reliance on abbreviations and greater potential for variability in terms of the clinical (and social, administrative or professional) statements included. The indications section is also completed by a far greater number of clinicians from various specialties whilst the findings section is limited to radiologists and sonographers.

In addition to high levels of accuracy, ToKSA offered substantial improvements in the efficiency of annotation with almost 50% of the reports receiving a label after only 2,000 sentences were annotated. Annotation of 3,407 full reports generated labels for only around 5% of the cohort yet each report contained multiple sentences and as such required a far greater reading time. The anticipated reading time to read all of the words in the full reports compared to the 2,000 sentences was between 176 and 401 times longer depending on the labelling task. Therefore, ToKSA offers a time-saving approach to annotation of free text electronic health records with no appreciable loss in accuracy. Furthermore, provision of an enlarged training set for the CNN resulted in higher accuracy of the CNN than when trained on data with labels derived from full report annotation. Many existing reports describe annotation of only a small number of reports for NLP training[2,4,24] and would benefit from the methodology described here.

We recommend that ToKSA is used to rapidly generate labels for a large corpus of reports on which CNN and/or other NLP techniques can be trained and evaluated. ToKSA can also be used as an adjunct to traditional annotation in which reports without a ToKSA-derived label are then selected for further annotation to capture infrequent sentence structures. We also recommend annotation of a smaller subset of full reports, on which the performance of both the ToKSA and CNN can be tested prior to publication. We provide an R script with which the sentence summarization aspects of ToKSA can be conducted.

We are not aware of other studies describing our methodology for facilitating manual annotation. Similar approaches have previously been used for automated annotation as well as specific steps within our methodology. One existing approach[25] relied on a labelled sequential pattern (LSP) to identify sentences containing follow-up requests within radiology reports. The LSP captured sequences containing time periods and follow-up instructions and was used to split data into sentences likely to contain a follow-up request and sentences without. These sentences were then annotated by clinician raters. This report did not, however, describe any of the word token harmonization or grouping of sentences we used to maximize report coverage and does not appear to have been prioritized for frequent sentences. The authors did not provide code for conducting this technique and the technique was specific to follow-up rather than generic to any pathology. Similarly, other authors[26] have described annotation of sentences but not with simplification, grouping and prioritization based on term-frequency matrices.

The strengths of this study lie in demonstrating the accuracy and consistency of our novel approach. We demonstrate very high levels of interobserver agreement for dual annotation of the reports and even greater levels of agreement between the techniques for annotation of report findings. We also describe robust methodology for data wrangling that minimizes the impact of typographical errors, accounts for presumed negation (e.g. “normal gallbladder” implying no gallstones unless otherwise described) and groups sentences with equivalent but non-identical terms to improve data quality and maximize the efficiency of annotation.

There are some limitations to consider. By purposefully focusing on annotating subunits of patient-level data common to several records, it is possible that NLP techniques trained on data with these labels will perform better for common report structures compared to records with less common structures. However, the performance of the CNN remained high when we considered only reports not annotated by ToKSA with accuracy of 98.3-100%. A hybrid approach with annotation of some full reports which do not receive an annotation from ToKSA may ensure some of the more unusual reports are also covered. ToKSA does not allow for inter-sentence co-reference meaning that the true meaning of some sentences cannot be determined without first reading the preceding sentence. This issue can be overcome by annotating any sentences in which inter-sentence co-reference is suspected and performing a second round of annotation with either the preceding sentence added or the full report. There is a possibility that some unknown term was used by a radiologist to refer to gallstones or the gallbladder. We took five steps to minimize this possibility. 1. We included structures in which gallstones are seen (the possibility of a radiologist describing gallstones by another synonym but without describing their location is implausible). 2. We confirmed in the 3,407 full reports that no terms-of-interest were missed. 3. We confirmed that no terms-of-interest were missed in the top 1,000 most frequently occurring sentences that did not contain a term-of-interest. 4. We searched the reports not flagged as containing a term-of-interest using regex. 5. We conducted a spell-check based on Damerau-Levenshtein distance [12–14] to ensure typographical errors did not conceal any terms-of-interest. Any remaining terms-of-interest were presumed to be extremely rare if present at all. Finally, our approach grouped sentences based on syntactic similarity and could be extended to incorporate semantic similarity, deduplication of synonyms and several other simplification steps prior to annotation. Extension of ToKSA, however, should be undertaken with caution if the approach is to remain generalizable to other diseases or phenotypes and we recommend that accuracy is always evaluated within a testing subset.

## CONCLUSION

In conclusion, we demonstrate a robust and efficient approach to maximising annotations of radiology reports with high levels of accuracy. Our approach enables annotation of tens of thousands of reports with human oversight, overcoming the obstacle of laborious report-by-report annotation and can be used as an adjunct to other NLP approaches.

## Data Availability

As data comprises confidential free text records from electronic healthcare records data cannot be made publicly available. A script used to analyse the data has been made available through a URL in the manuscript.

https://github.com/SurgicalInformatics/ToKSA

## ACKNOWLEDGEMENTS AND FUNDING

This work was funded by a Medical Research Council Clinical Research Training Fellowship awarded to CJF (MR/T008008/1). NLM is supported by the British Heart Foundation through a Chair Award (CH/F/21/90010) and a Research Excellence Award (RE/18/5/34216).

## AUTHOR CONTRIBUTIONS

CJF prepared the draft manuscript, analyzed the data, developed the ToKSA methodology and applied for ethical approval.

EMH, AS and SJW conceptualized the project and assisted with cohort establishment.

CJF, WC, LC and SRK annotated the data.

TMD, RP, EMH and CAS assisted with project administration, server maintenance and software installations

NM, EMH and CJF were involved in data acquisition and storage

HW and NLM contributed to refinement of the methodology

All authors reviewed the manuscript and provided feedback on the methods

