## Supplementary Materials for "ToKSA - Tokenized Key Sentence Annotation - a Novel Method for Rapid Approximation of Ground Truth for Natural Language Processing"

Supplementary Materials: Methods, Tables and Figures

### Supplementary Methods

#### Regex and Term-specific Glossary

Regex was used to select relevant sentences most likely to contain the information necessary to complete annotation and generate a label. The regex was used to “tag” sentences with terms of interest that were then included in the 1,000 most frequent sentences with a term of interest. The additional 1,000 most frequent sentences without a term of interest were inspected to ensure that no relevant terms were excluded despite being commonly used.

The regex used to identify terms of interest in the “indication” section was designed to cover symptoms, blood results, syndromes and structures related to gallstone pathology:

‘calcul|stone|lith|cholecyst|gallbladder|\\bgb\\b|biliary|ruq|lft|amylase|pancreat|mir(r)?iz(z)?i|bouv(e)?ret’

The regex to identify terms of interest in the “findings” was designed to include commonly used radiological terms relevant to gallstone pathology and relevant hepatobiliary structures:

‘stone|calcul|sludge|debris|grit|(chole|choledocho|hepato|pancreato)[ -]*lithiasis|chole[ -]*cyst|gall|biliary’

The same regex was used for both gallstone status and cholecystectomy status as the pathology and anatomical structures were closely related. Use of anatomical structures ensured that rare or misspelled disease terms were likely to be identified through the anatomical structure, furthermore “normal gallbladder and biliary tree” can be interpreted as the absence of gallstones despite not including a disease term. After identification of sentences containing a term of interest from the “findings” section, sentences containing the bigrams (two-word N-grams) “kidney”, "ureter*" or “bladder” - “stone” or “calculus” were excluded (the term “bladder” was not dropped if “gall” also formed a trigram with “bladder” and “stone” e.g. “gall bladder stone”).

#### Annotation for Symptom, Gallstone and Cholecystectomy Status

For the indication section the sentences were classified in the following way:

1. Symptom status:
   - Symptoms suspicious for gallstones
     - Abdominal pain
     - Jaundice or deranged liver function
     - Prior history of or high clinical suspicion of gallstones or gallstone-related pathology
   - Not suspicious for gallstones
     - Clear alternative pathology (e.g. hepatocellular carcinoma screening)
     - Symptoms unrelated to gallstones (e.g. pulsatile mass, trauma, hematuria, etc.)

For the findings section the sentences were classified in the following way:

2. Presence of gallstones
   - Gallstones definitely seen
   - Gallstones excluded for a patient (e.g. “Conclusion: No gallstones.”)
   - Gallstones excluded but only for specific location (e.g. “Normal gallbladder.” - this does not fully exclude a separate sentence in the same report indicating gallstones elsewhere in the biliary tree)
   - No mention of gallstones or related structures, or unable to provide definitive statement regarding gallstones
3. Presence of cholecystectomy
   - Cholecystectomy confirmed
   - No definitive evidence of cholecystectomy or gallbladder seen (these were labelled separately before being combined into one label)

#### ToKSA Process for Ultrasound Reports

1. Text was converted to lower case as capitalization will not change the interpretation for the purposes of this study
2. The "indication" and "findings" sections were tokenized into single words and a term-frequency matrix was generated (this is a matrix of all words and the number of times they occur in the entire corpus of reports)
3. A term-specific glossary (for the "indication" section: symptoms relevant to the disease and disease terms; for the "findings" section: disease terms and anatomic structures affected by the disease) was created and supplemented with regex to account for subtle variations in spelling, punctuation and word-spacing
4. Spelling mistakes were accounted for using the Damerau-Levenshtein distance (edit distance allowing for typographical errors including substitutions, insertions, deletions and transpositions
   - Each word in the term-frequency matrix was joined with any terms of interest identified using the edit distance
   - This was performed only for the term-specific glossary and using an edit distance of 2 or less
   - Terms with a string length of 4 or less (e.g. "grit") were limited to an edit distance of 1 to minimize excessive matching on dissimilar small strings
5. All misspelled terms of interest were overwritten with the correct spelling after a manual inspection
   - Terms which incorrectly matched (e.g. "debride" which is an edit distance of 2 from "debris") were not overwritten
   - This approach resulted in corrected spelling of any relevant words from the term-specific glossary whilst remaining reliant on manual adjudication
6. Further searching with regex was conducted to identify and overwrite misspellings with an edit distance greater than 2 - for instance "gallstonesseen" (an edit distance of 4 from "gallstones" but evidently missing a single space)
7. Regex was used to overwrite patterns which may emerge in the reports that are not relevant to the research question and increase the number of unique sentences
   - The matched patterns included patient identification number, telephone number, physician registration number, email address, dates, year and measurements (e.g. "12mm")
   - Words were overwritten by the type of matched pattern (e.g. "2007" was converted to "[year]")
   - This ensured that sentence meaning was not altered by removal of these terms
8. A new term-frequency matrix was generated encompassing word substitutions from steps 7-9
9. Each term occurring 5 or fewer times was overwritten with "[redacted]"
10. The reports were then tokenized into whole sentence N-grams
11. The sentences were arranged into a term-frequency matrix and sorted based on the frequency of the occurrence of the sentence
12. Two groups of sentences were taken forward for annotation: the 1000 most frequent sentences which contained a term of interest (from the term-specific glossary) and the 1000 most frequent sentences without a term of interest (the latter was inspected to ensure no disease terms or anatomical structures were missed from the term-specific glossary).
13. The sentences were annotated with desired labels
14. The labels generated for each sentence were combined into full report labels (a single sentence confirming the presence of disease is sufficient to generate a positive disease label for the full report; when all sentences in the report containing a term-of-interest describe the absence of the disease then a negative disease label can be applied to the full report)
15. Any conflicting labels were resolved through inspection of the full report (in theory, no reports should receive contradictory classifications e.g. both “definite gallstones” and “no gallstones”, and such dual classifications were sought and if present assessed for the reason underlying this)

#### Combination of Sentence Labels

After annotation of the most frequent sentences, the generated labels were combined into a single label per report. Some reports had 1 sentence with a label, some had none and some had multiple.

For the “indication” section the labels were combined in the following steps:

- Any reports with a label in which one or more labels were “Suspicious” for gallstones (e.g. “right upper quadrant pain.”) were considered as “Suspicious” regardless of other sentences in the same report
- Reports which only contained sentences with a label of “Not suspicious” for gallstones which had no sentences without a label (i.e. all sentences from the report were within the 2,000 annotated) were labelled as “Not suspicious”
- Reports which contained at least one sentence with a label of “Not suspicious” for gallstones, had no sentences with a label of “Suspicious” for gallstones and no sentences with a term of interest that lacked a label were also classified as “Not suspicious”
- Reports which contained at least one sentence with a label of “Not suspicious” for gallstones but contained sentences with a term of interest that lacked a label did not receive a label from ToKSA as the remaining sentences may include symptoms “Suspicious” for gallstones
- Reports that contained no sentences with a label did not receive any combined label

For the “findings” section gallstone status labels were combined in the following steps:

- Any reports which contained a sentence with a label of “Gallstones” were considered as “Gallstones” regardless of other sentences within the report
- Reports containing sentences thought to definitively rule out stones (e.g. “opinion: no gallstones seen on scan.”) received a combined label of “No gallstones”
- Reports in which all sentences containing a term of interest received a label of “No gallstones” even if only within specified organs (e.g. “No stones seen in gallbladder”) were given a label of “No gallstones”
- Reports which contained any sentences with a term of interest that did not receive a label from ToKSA and did not have a sentence confirming gallstones did not receive a combined label (one sentence with a label such as “no stones were seen in the gallbladder.” may in theory be followed by “a solitary stone is seen in the common bile duct”, therefore combining negative labels was not possible in the situation of relevant sentences without a label)

For the “findings” section cholecystectomy status labels were combined in the following steps:

- Any reports which contained a sentence with a label of “Cholecystectomy” were considered as “Cholecystectomy” regardless of other sentences within the report
- Reports which contained a sentence with a label of “Gallbladder seen” were considered as “No evidence of cholecystectomy” regardless of other sentences within the report
- Reports in which all sentences containing a term of interest received a label of “No evidence of cholecystectomy” or “Gallbladder seen” were given a label of “No evidence of cholecystectomy”
- Reports which contained any sentences with a term of interest that did not receive a label from ToKSA and did not have a sentence confirming cholecystectomy or the presence of a gallbladder did not receive a combined label as cholecystectomy status remained unclear

### Supplementary Tables

| **Supplementary Table 1: Frequent Sentences “Indication” Section** | | |
| --- | --- | --- |
| **Sentence** | **Number of times seen** | **Label** |
| query gallstone | 5818 | Suspicious |
| deranged lfts | 2747 | Suspicious |
| abnormal lfts | 2152 | Suspicious |
| ruq pain | 1549 | Suspicious |
| query cholecystitis | 1214 | Suspicious |
| query biliary colic | 772 | Suspicious |
| previous cholecystectomy | 597 | Suspicious |
| tender ruq | 513 | Suspicious |
| query gall stone | 429 | Suspicious |
| lfts normal | 422 | Not Suspicious |
| known gallstone | 331 | Suspicious |
| deranged liver enzymes | 325 | Suspicious |
| normal lfts | 316 | Not Suspicious |
| newly deranged lfts | 298 | Suspicious |
| jaundice | 271 | Suspicious |
| query gallstone pancreatitis | 233 | Suspicious |
| lfts deranged | 209 | Suspicious |
| query acute cholecystitis | 208 | Suspicious |
| query gallstone / ultrasound abdomen | 205 | Suspicious |
| query biliary pathology | 195 | Suspicious |

| **Supplementary Table 2: Frequent Sentences “Findings” Section** | | | |
| --- | --- | --- | --- |
| **Sentence** | **Number of times seen** | **Label (Gallstones)** | **Label (Cholecystectomy)** |
| no hepato-biliary dilatation demonstrated | 12712 | Unknown | No evidence of cholecystectomy |
| no biliary dilatation | 11265 | Unknown | No evidence of cholecystectomy |
| normal appearances gallbladder and cbd | 7432 | No Gallstones (possible) | Gallbladder seen |
| no gallstone | 4798 | No Gallstones (definite) | No evidence of cholecystectomy |
| the biliary tree is not dilated | 4192 | Unknown | No evidence of cholecystectomy |
| no biliary tree dilatation | 4166 | Unknown | No evidence of cholecystectomy |
| biliary system was not dilated | 4119 | Unknown | No evidence of cholecystectomy |
| cholecystectomy noted | 3253 | Unknown | Cholecystectomy |
| no dilatation of the biliary tree | 2255 | Unknown | No evidence of cholecystectomy |
| the gallbladder was normal with no calculus | 2197 | No Gallstones (possible) | Gallbladder seen |
| no intrahepatic biliary dilatation | 1946 | Unknown | No evidence of cholecystectomy |
| the gallbladder appears normal | 1906 | No Gallstones (possible) | Gallbladder seen |
| normal gallbladder and biliary tree | 1892 | No Gallstones (possible) | Gallbladder seen |
| the gallbladder and biliary tree appear normal | 1786 | No Gallstones (possible) | Gallbladder seen |
| normal gallbladder | 1732 | No Gallstones (possible) | Gallbladder seen |
| no intra or extra hepatic biliary dilatation | 1571 | Unknown | No evidence of cholecystectomy |
| no intra or extrahepatic biliary dilatation | 1516 | Unknown | No evidence of cholecystectomy |
| previous cholecystectomy | 1412 | Unknown | Cholecystectomy |
| no pericholecystic fluid | 1382 | Unknown | Gallbladder seen |
| the gallbladder, cbd and biliary tree appear normal with no intrahepatic duct dilatation or stone seen | 1350 | No Gallstones (definite) | Gallbladder seen |
| *As discussed in Supplementary Methods – ToKSA generated some definite gallstone labels which were used to provide definite label regardless of remaining report. Some were considered possible and final label depended on the presence of other sentences within the report. | | | |

| **Supplementary Table 3: Interobserver Agreement between Human Raters** | | |
| --- | --- | --- |
| Classification | Weighted Cohen’s Kappa | Weighted Bangdiwala’s B-Statistic |
| Indication (Whole Report) | 0.992 | 0.995 |
| Findings-Gallstones (Whole Report) | 0.956 | 0.993 |
| Findings-Cholecystectomy (Whole Report) | 1.000 | 1.000 |
| Indication (Sentences Only) | 0.965 | 0.965 |
| Findings-Gallstones (Sentences Only) | 0.985 | 0.997 |
| Findings-Cholecystectomy (Sentences Only) | 0.997 | 0.999 |

| **Supplementary Table 4: Confusion Matrix of ToKSA versus Full Report Labels**  **(Symptom Status)** | | |
| --- | --- | --- |
|  | Tokenized Sentence Annotation Label | |
| Full Report Label | Suspicious for gallstones | Not suspicious for gallstones |
| Suspicious for gallstones | 1572 | 12 |
| Not suspicious for gallstones | 17 | 212 |

| **Supplementary Table 5: Confusion Matrix of ToKSA versus Full Report Labels**  **(Gallstone Status)** | | | |
| --- | --- | --- | --- |
|  | Tokenized Sentence Annotation Label | | |
| Full Report Label | Gallstones | No Gallstones | Unknown |
| Gallstones | 315 | 2 | 0 |
| No Gallstones | 0 | 1723 | 14 |
| Unknown | 0 | 0 | 3 |

| **Supplementary Table 6: Confusion Matrix of ToKSA versus Full Report Labels**  **(Cholecystectomy Status)** | | |
| --- | --- | --- |
|  | Tokenized Sentence Annotation Label | |
| Full Report Label | Cholecystectomy | No evidence of cholecystectomy |
| Cholecystectomy | 34 | 0 |
| No evidence of cholecystectomy | 0 | 1639 |

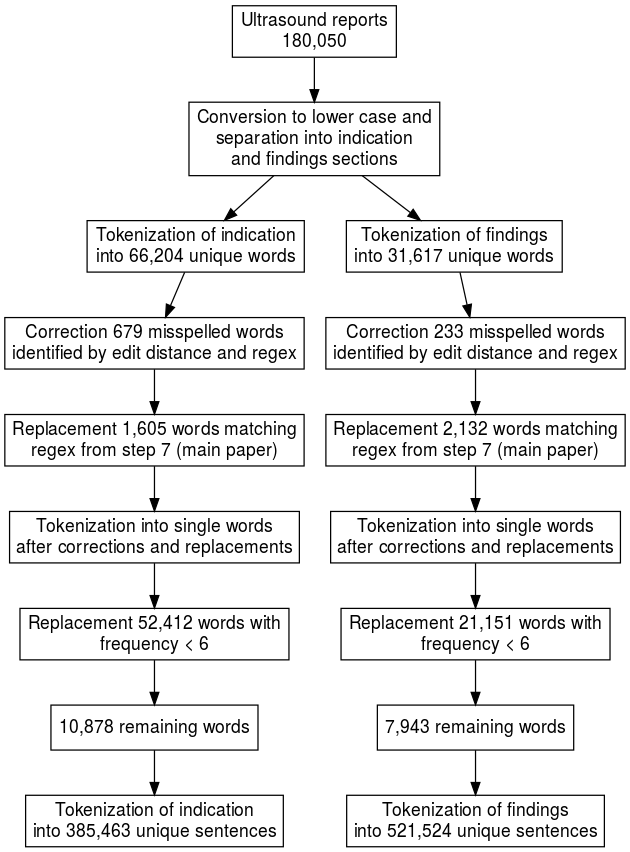
**Supplementary Figure 1**. Flowchart showing tokenization and text cleaning undertaken during Tokenized Key Sentence Annotation
